# A non-specialist worker delivered digital assessment of cognitive development (DEEP) in young children: a longitudinal validation study in rural India

**DOI:** 10.1101/2024.11.04.24316724

**Authors:** Supriya Bhavnani, Alok Ranjan, Debarati Mukherjee, Gauri Divan, Amit Prakash, Astha Yadav, Chaman Lal, Diksha Gajria, Hiba Irfan, Kamal Kant Sharma, Smita Dattatraya Todkar, Vikram Patel, Gareth McCray

**Author notes:** Corresponding author; Prof Vikram Patel, Paul Farmer Professor and Chair, Department of Global Health and Social Medicine, Harvard Medical School. joint first authors (Supriya Bhavnani, Alok Ranjan). joint senior authors (Vikram Patel, Gareth McCray).

## Abstract

**Background:** Cognitive development in early childhood is critical for life-long well-being. Existing cognitive development surveillance tools require lengthy parental interviews and observations of children. Developmental Assessment on an E-Platform (DEEP) is a digital tool designed to address this gap by providing a gamified, direct assessment of cognition in young children which can be delivered by front-line providers in community settings.

**Methods:** This longitudinal study recruited children from the SPRING trial in rural Haryana, India. DEEP was administered at 39 (SD 1; N=1359), 60 (SD 5; N=1234) and 95 (SD 4; N=600) months and scores were derived using item response theory. Criterion validity was examined by correlating DEEP-score with age, Bayley’s Scales of Infant Development (BSID-III) cognitive domain score at age 3 and Raven’s Coloured Progressive Matrices (CPM) at age 8; predictive validity was examined by correlating DEEP-scores at preschool-age with academic performance at age 8 and convergent validity through correlations with height-for-age z-scores (HAZ) and early life adversities.

**Findings:** DEEP-score correlated strongly with age (r=0.83, 95% CI 0.82-0.84) and moderately with BSID-III (r=0.50, 0.39-0.60) and CPM (r=0.37; 0.30 – 0.44). DEEP-score at preschool-age predicted academic outcomes at school-age (0.32; 0.25 – 0.41) and correlated positively with HAZ and negatively with early life adversities.

**Interpretation:** DEEP provides a valid, scalable method for cognitive assessment. It’s integration into developmental surveillance programs could aid in monitoring and early detection of cognitive delays, enabling timely interventions.

**Funding:** SPRING, REACH and COINCIDE were funded through Wellcome Trust, Madura Microfinance Ltd and Wellcome Trust/DBT India Alliance respectively.

## Introduction

Numerous longitudinal studies have demonstrated that cognitive development in early childhood is predictive of schooling attainment, mental health and adult Intelligence Quotient (IQ),^1,2^ and is thus critical to the well-being and economic productivity of individuals across their life course.^3^ The fact that the highest rate of economic returns comes from investing in this period, illustrated by Heckman’s curve,^4^ is recognised globally, including in the Sustainable Development Goals. In India too, the National Education Policy announced in 2020 has listed “the highest priority to achieving foundational literacy and numeracy by all students by Grade 3 (8-years age)” within its fundamental principles. Despite the knowledge of the importance of the preschool years, millions of children in low- and middle-income countries (LMICs), including India, have sub-optimal cognitive development through this period resulting in poor school readiness.^5^ This is largely due to a disproportionately high burden of early-life adversities in LMICs, often associated with poverty, contributing to the lack of nurturing and safe environments which are essential for healthy brain development, and which lead to a vicious cycle of intergenerational transmission of disadvantage.^6^ The recent Annual Status of Education Report (ASER) of India observed that less than a quarter of children in primary school are at expected level for reading and math.^7^ Additionally, the estimated prevalence of intellectual disability ranges from 3.1% in children aged 2-6 to 5.2% in children aged 6-9 years, translating to tens of millions of children in need of support.^8^ Thus, assessment and monitoring of cognitive development in early childhood, when the brain is most plastic and sensitive to interventions, to identify delays and disruptions in a timely manner, is essential to institute early interventions to promote cognitive development and improve educational and mental health outcomes across the life course.

However, many children who would benefit most from early interventions do not get identified in a timely manner. A major barrier to identifying children with delayed or disrupted brain development is low awareness of age-appropriate developmental milestones in communities and the lack of routine cognitive developmental surveillance, analogous to growth monitoring.^9^ This results in children being typically detected later in childhood, often when they experience educational difficulties, and well after the critical sensitive period for early interventions has passed. Furthermore, cognitive development assessments rely on expensive, proprietary, time-intensive, observational tools which can only be administered by highly trained, and scarce, child development specialists. Governments of LMICs are making concerted efforts to overcome the barrier of the lack of specialist providers by employing the strategy of task-sharing,^10^ in which front-line workers are trained to perform a range of tasks dedicated to the health and well-being of infants and children. Efforts have recently been made to develop and validate globally relevant open-source tools for the assessment of child development which can be used by such front-line workers, such as the Caregiver Reported Early Development Instruments (CREDI),^11^ Early Childhood Development Index (ECDI),^12^ Global Scales for Early Development (GSED)^13^ and the International Development and Early Learning Assessment (IDELA).^14^ However, these tools rely either on the subjective responses of parents to questionnaires or on behavioural observations made by front-line workers, both of which may introduce biases. These limitations can be addressed by measures which directly assess child performance and, thereby, do not rely on potentially unreliable administrator judgement or parent report.

The Developmental Assessment on an E-Platform (DEEP), is an Android tablet-based tool for measuring cognition in children aged 2.5-6-years. It comprises a battery of 14 games, developed from tasks used in clinical developmental assessments, with most games having multiple levels of difficulty. Nine of these games were created to measure cognition in 3-year-old children and have been previously described.^15^ Five were subsequently added for cognition in older children (up to 6-years). These games measure a range of cognitive constructs including reasoning, response inhibition, categorisation, memory and visual form perception and integration. DEEP was designed for scalability in several ways: it is delivered on routinely available tablet devices; it is administered by non-specialist workers; it does not require fluency in any specific language; and it does not require an internet connection for completion of the assessment. Studies have found DEEP to be highly acceptable to 3-year-old children, as indicated by high completion rates.^15^ A proof-of-concept paper has demonstrated that DEEP-scores can be derived from metrics of children’s performance on the games by using supervised machine learning benchmarked to the gold-standard cognitive assessment, Bayley’s Scale of Infant and Toddler Development – 3^rd^ Edition.^16,17^ However, the proof-of-concept was limited by the small sample size of the training and test datasets and the cross-sectional data. The present study applies the principles of item response theory (IRT) to generate DEEP-scores based on psychometric principles. IRT is a psychometric framework to transparently model the relationship between responses to items or other metrics and unobserved latent traits.^18^ It has over half-a-century of use in general educational settings, but has only recently been applied to measure development in early childhood.^19,20^ The aim of this paper is to assess the criterion, predictive and convergent (equivalent to ‘hypothesis testing’ in COSMIN checklist^21^) validity of DEEP-score, to generate evidence for its utility as a scalable cognitive assessment for preschool children.

## Methods

### Study population

The SPRING cluster randomized controlled trial recruited children born on or after June 2015 from 120 villages in Rewari district of rural Haryana, India.^22^ Seven-thousand-and-fifteen families were enrolled by the surveillance system from 24 clusters, defined as the catchment area of a functional primary health sub-centre. Trial outcome measures were assessed in 1443 children at 18 months of age by the SPRING study, which formed the sampling frame of this study.

One-thousand-three-hundred-and-fifty-nine children were enrolled into this study at 3-years age (BL) and have been followed-up 2 times through the completed REACH and ongoing COINCIDE studies (Figure 1).^23^ For FU1, 1304 children from the SPRING outcome cohort were followed up between December 2019 and April 2021 when they were 4-6-years old. Data was collected from 1234 children and 70 were lost to follow-up. Finally, through the COINCIDE study, data was collected from 600 children (reduced sample size due to funding limitations of the COINCIDE study) when they were approximately 8-years-old (FU2). This sample were purposively selected to ensure their socio-demographic characteristics were comparable to FU1 and FU2.

**Figure 1:**
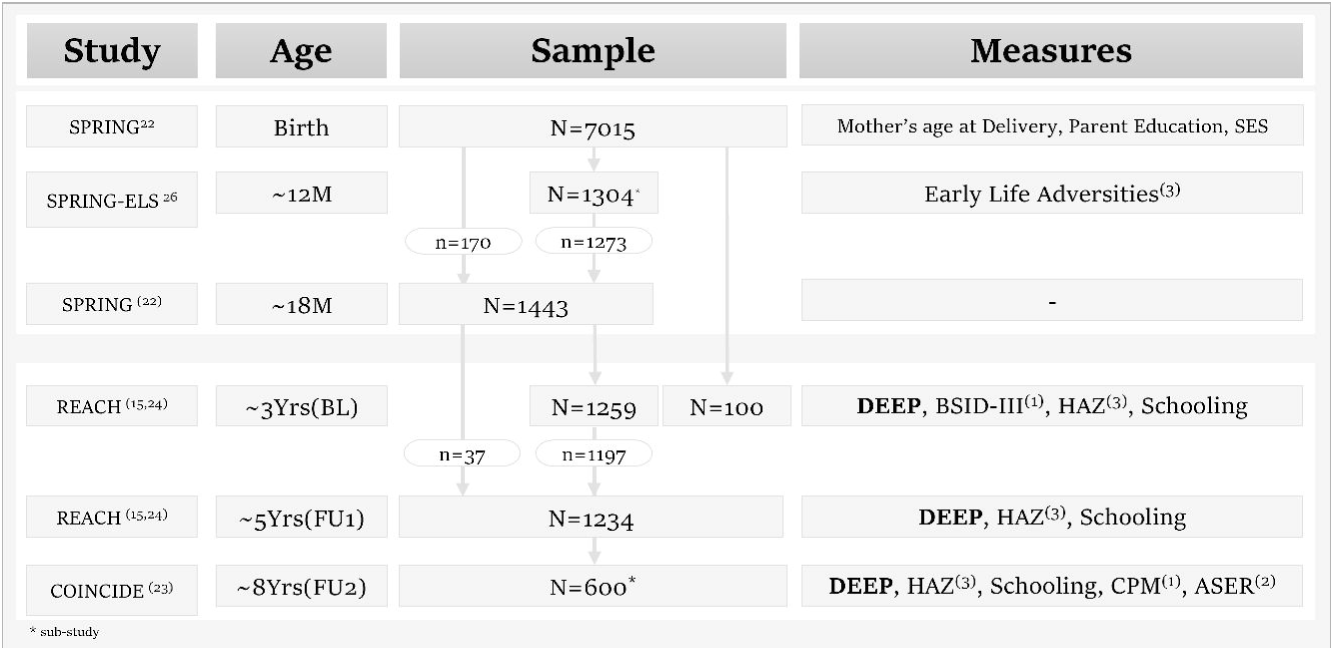
A flowchart of the participants of SPRING, REACH and COINCIDE studies and measures used in this study to evaluate the criterion (1), predictive (2) and convergent (3) validity of the DEEP tool. .

Written informed consent was taken from parents, and verbal assent from children in FU2, for participation in this study. Ethical approval for the studies which collected the data reported in this paper was obtained from Sangath’s Institutional Review Board (GD_2019_55, 28 August 2019 and GD_2022_77, 4 August 2022).

### Data collection

The assessments on 3-year-old children (BL) have been previously described^24^ and all assessments in FU1 and FU2 were conducted in a similar manner by non-specialists (henceforth referred to as ‘assessors’) in participants’ households at a convenient date and time. These assessors had completed the equivalent of a postgraduate degree, were embedded within the community through prior work and had training and experience working with young children. Data was collected either on a Huawei MediaPad T5 tablet (BL and FU1) or a Samsung Galaxy Tab A8 tablet (FU2). Approximately 10% of all visits were overseen by a field supervisor, who was closely supported by senior researcher team members, to ensure fidelity to administration protocols. Weekly group meetings between the field supervisor and all assessors were used to provide peer support and regular feedback, and quarterly refresher trainings were conducted by senior research team members.

### Measures

#### Developmental Assessment on an E-Platform (DEEP)

The 14 games comprising the DEEP tool are described in Table 1. At BL, 3-year-old children played only 9 games while at FU1 and FU2, children played the larger suite of 14 games, including the original 9 games - some with an increased number of difficulty levels. Children interact with DEEP through the use of *tap* or *drag and drop* gestures. The main cognitive constructs which each game targets are listed, but it is expected that each game taps into multiple constructs and thus that each construct is represented in more than one game. The following metrics were derived from each DEEP game level (See Supplementary Materials for details): Accuracy - Proportion of correct clicks; Completion_time - Proportion of maximum time taken to complete the level; Latency - Time till first click or drag; Activity - Number of clicks or drags per second; and Highest_level - the number of difficulty levels played for each game. Modelling was done using the dataset from BL and FU1, since that represents the age for which DEEP has been created (2.5-6-year-olds), and the model was jointly fit to harmonise scores across the age groups making them interpretable on the same scale. The final model was used to derive DEEP-score for 8-year-olds (FU2). Graded response polytomous IRT models^25^ were fitted using maximum likelihood item factor analysis, in the *mirt* package in the R statistical software. Models were assessed based on a) root mean square error of estimation (RMSEA), b) Tucker-Lewis Index (TLI) and Comparative Fit Index (CFI), and more informally c) correlation with age (Supplementary Table 1). Subject matter experts made the final decision about which model to select based on expert knowledge and model fit statistics. The final model chosen included Accuracy and Completion_time. The discrimination, which indicates an item’s ability to differentiate between individuals with different levels of the underlying trait, and difficulty, which indicates the level of ability required to have a 50% chance of answering an item correctly, of test items were derived for the final model (Supplementary Table 2). Test reliability, and Standard Error of Estimation (SEE), which provides an estimate of the amount of error inherent in an individual’s observed score due to the imprecision of the tool, was also derived (Supplementary Figure 1).

**Table 1:**
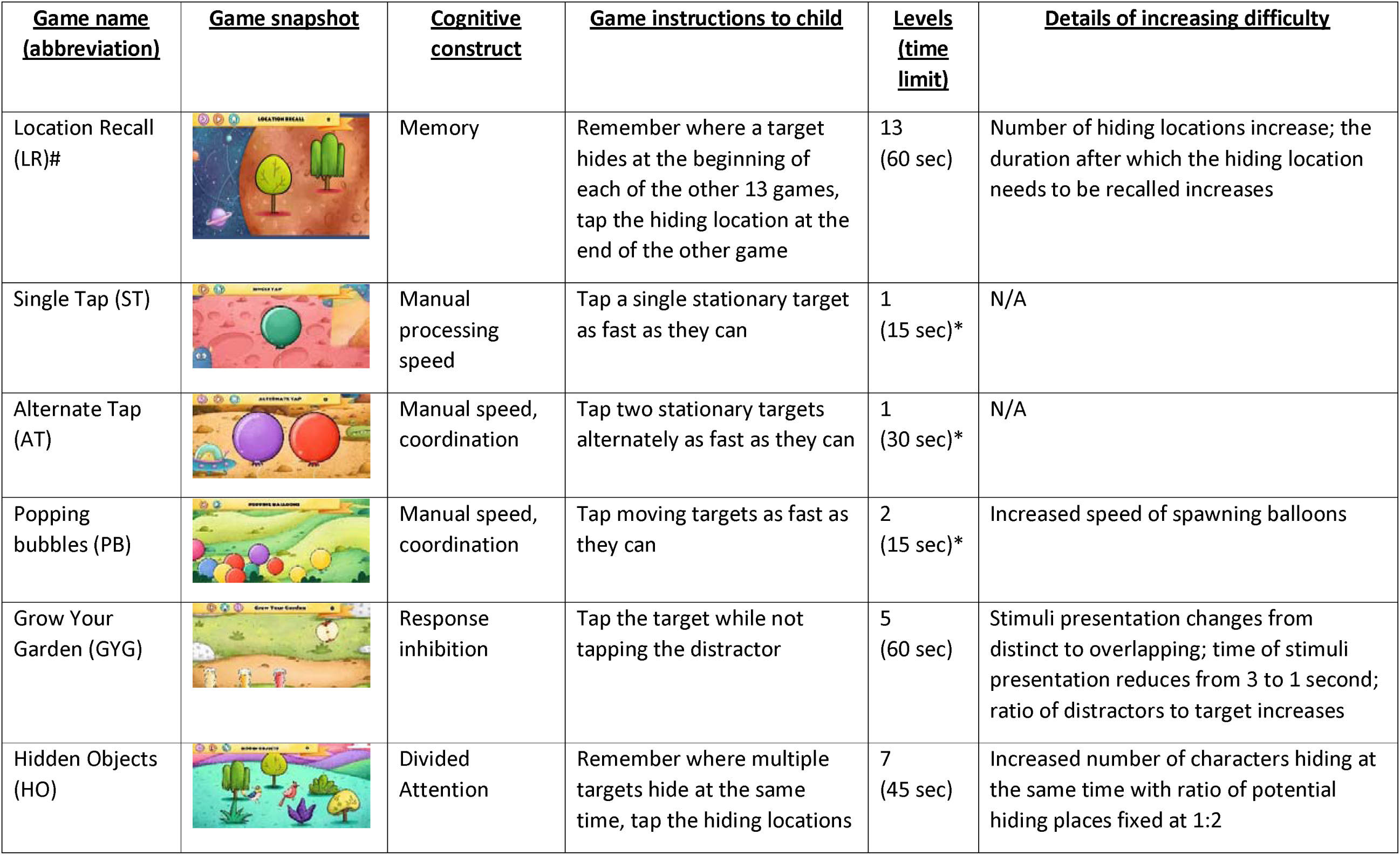
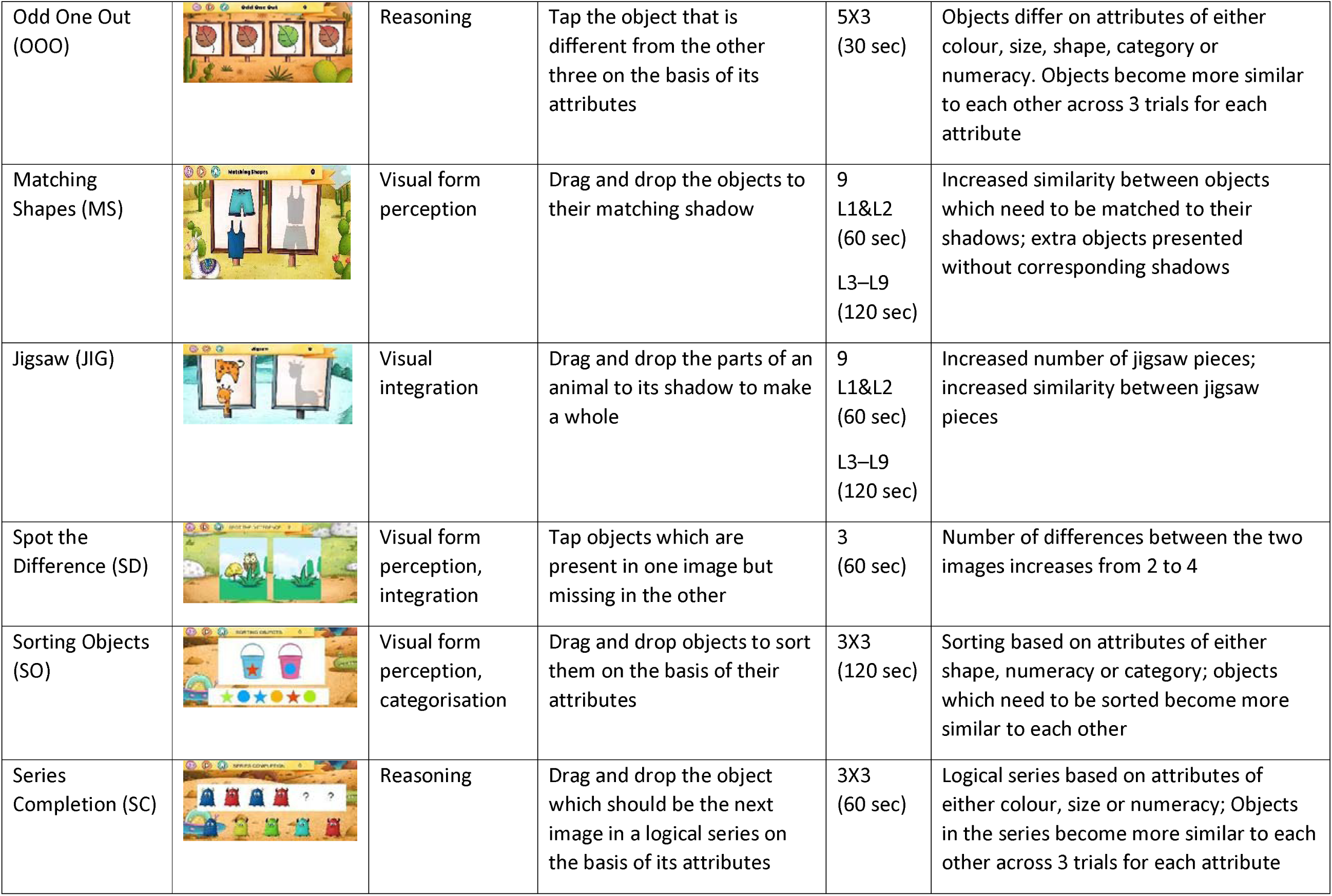
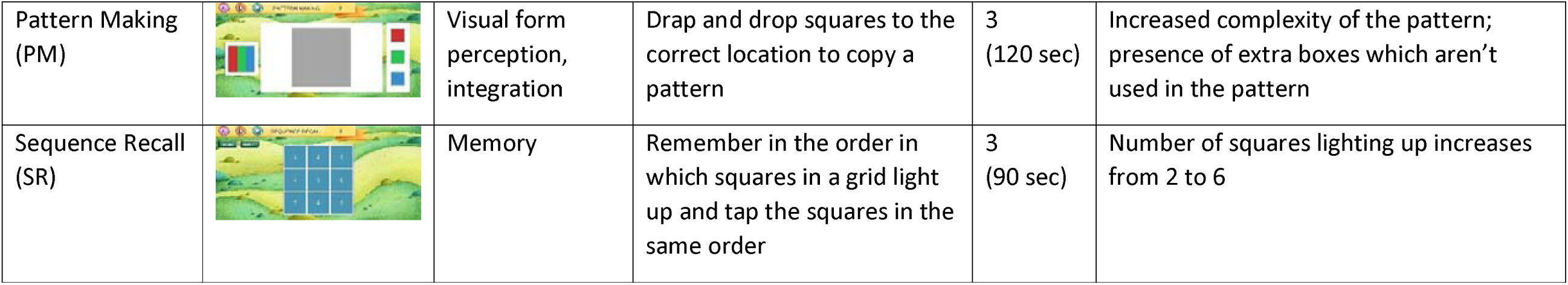
The DEEP tool games and their cognitive constructs.

| **<u>Game name</u>**<br>**<u>(abbreviation)</u>** | **<u>Game snapshot</u>** | **<u>Cognitive</u>**<br>**<u>construct</u>** | **<u>Game instructions to child</u>** | **<u>Levels</u>**<br>**<u>(time</u>**<br>**<u>limit)</u>** | **<u>Details of increasing difficulty</u>** |
| --- | --- | --- | --- | --- | --- |
| Location Recall (LR)# |  | Memory | Remember where a target hides at the beginning of each of the other 13 games, tap the hiding location at the end of the other game | 13<br>(60 sec) | Number of hiding locations increase; the duration after which the hiding location needs to be recalled increases |
| Single Tap (ST) |  | Manual processing speed | Tap a single stationary target as fast as they can | 1<br>(15 sec)* | N/A |
| Alternate Tap (AT) |  | Manual speed, coordination | Tap two stationary targets alternately as fast as they can | 1<br>(30 sec)* | N/A |
| Popping bubbles (PB) |  | Manual speed, coordination | Tap moving targets as fast as they can | 2<br>(15 sec)* | Increased speed of spawning balloons |
| Grow Your Garden (GYG) |  | Response inhibition | Tap the target while not tapping the distractor | 5<br>(60 sec) | Stimuli presentation changes from distinct to overlapping; time of stimuli presentation reduces from 3 to 1 second; ratio of distractors to target increases |
| Hidden Objects (HO) |  | Divided Attention | Remember where multiple targets hide at the same time, tap the hiding locations | 7<br>(45 sec) | Increased number of characters hiding at the same time with ratio of potential hiding places fixed at 1:2 |
| Odd One Out (OOO) |  | Reasoning | Tap the object that is different from the other three on the basis of its attributes | 5X3<br>(30 sec) | Objects differ on attributes of either colour, size, shape, category or numeracy. Objects become more similar to each other across 3 trials for each attribute |
| Matching Shapes (MS) |  | Visual form perception | Drag and drop the objects to their matching shadow | 9<br>L1&L2<br>(60 sec)<br>L3-L9<br>(120 sec) | Increased similarity between objects which need to be matched to their shadows; extra objects presented without corresponding shadows |
| Jigsaw (JIG) |  | Visual integration | Drag and drop the parts of an animal to its shadow to make a whole | 9<br>L1&L2<br>(60 sec)<br>L3-L9<br>(120 sec) | Increased number of jigsaw pieces; increased similarity between jigsaw pieces |
| Spot the Difference (SD) |  | Visual form perception, integration | Tap objects which are present in one image but missing in the other | 3<br>(60 sec) | Number of differences between the two images increases from 2 to 4 |
| Sorting Objects (SO) |  | Visual form perception, categorisation | Drag and drop objects to sort them on the basis of their attributes | 3X3<br>(120 sec) | Sorting based on attributes of either shape, numeracy or category; objects which need to be sorted become more similar to each other |
| Series Completion (SC) |  | Reasoning | Drag and drop the object which should be the next image in a logical series on the basis of its attributes | 3X3<br>(60 sec) | Logical series based on attributes of either colour, size or numeracy; Objects in the series become more similar to each other across 3 trials for each attribute |
| Pattern Making<br>(PM) |  | Visual form<br>perception,<br>integration | Drag and drop squares to the<br>correct location to copy a<br>pattern | 3<br>(120 sec) | Increased complexity of the pattern;<br>presence of extra boxes which aren't<br>used in the pattern |
| Sequence Recall<br>(SR) |  | Memory | Remember in the order in<br>which squares in a grid light<br>up and tap the squares in the<br>same order | 3<br>(90 sec) | Number of squares lighting up increases<br>from 2 to 6 |

#### BSID-3^rd^ edition

The Bayley’s Scale of Infant and Toddler Development, 3rd Edition (BSID-III), a developmental assessment for preschool children aged 0-42 months^16^, was administered on a subset of 200 children at BL. A translated version of the BSID-III adapted for administration by non-specialists was used following a protocol described previously^24,26^. Raw scores were computed as per the manual and used to generate age-adjusted composite scores.

#### Raven’s Coloured Progressive Matrices (CPM)

CPM was administered in FU2, when children were 8-years-old. CPM measures fluid intelligence and non-verbal reasoning abilities of 5-11-year-old children, and comprises three sets of 12 items each of increasing difficulty. Set A measures predominantly visuoperceptual abilities, Set Ab configuration processing and Set B mainly analogical reasoning. It has extensively been used in the Indian population and Indian norms were used to create age-adjusted standardised scores.

#### Annual Status of Education Report (ASER) tool

Literacy and numeracy was assessed in FU2 using the ASER tool that has previously been used in India. Stimuli are presented using flip books and items are comparable to widely used tools such as the Early Grade Reading Assessment (EGRA). *Early life adversity:* Details of how early life adversity was measured and computed in the SPRING study is described in detail elsewhere.^26^ Twenty-two contextually relevant adversities were selected and categorized into four domains: socio-economic factors (SES), maternal stress, quality of relationships of the child with their caregivers and finally, direct stressors to the child. A sum of all adversities experienced by the child was derived to represent their cumulative adversity.

#### Anthropometry

World Health Organisation (WHO) protocols were used to measure the child’s height using the Seca 213 Portable Stadiometer and height-for-age (HAZ) z-scores were generated using WHO growth standards. Stunting was defined as two standard deviations below the age-adjusted WHO growth-standard median values of height. All children whose age-adjusted anthropometric measurements were below three standard deviations of WHO median values were referred for follow-up assessments to local clinics.

#### Socio-demographic information

Data on parental education and socioeconomic status was collected from families at enrolment through the SPRING study.^22^ Principal components analysis (PCA) was used to calculate a socioeconomic status (SES) index using data on household demographics and animal & other asset ownership. This index was used to categorize the population into SES quintiles.

### Statistical analyses

All relationships between DEEP-score and validity measures have been described using Pearson’s correlation coefficients with 95% confidence intervals (CIs). Criterion Validity is examined through comparison with chronological age across the range of 2.5-8-years and concurrently administered BSID-III at BL and CPM at FU2; Predictive Validity is examined through prediction of ASER scores by DEEP-score at BL and FU1; Associations of DEEP-score at BL, FU1 and FU2 with concurrently measured HAZ and with early life adversities provides evidence of Convergent Validity. Note, a sensitivity analysis is presented for the cumulative adversity score with and without the relationship domain of early life adversities, which had missing data for 32.8% children in the SPRING study.^26^ Statistically significant p values are represented as asterisks. All analyses were conducted using R 4.2.1.

## Results

### Description of study participants

The socio-demographic details of study participants at BL and follow-ups 1 and 2 are described and compared to the entire database of children enrolled in the SPRING study in Table 2. Mean age of children at BL was 39 months (SD: 1), 45.9% were female and 44.7% attended preschool. At FU1, mean age of children was 60 months (SD: 5), 45.7% were female and most children (91.4%) attended preschool. At FU2, mean age of children was 95 months (SD: 4), 45.2% were female and all children attended either public or private school. A majority of caregivers (59-63.5% mothers and 72.8-77.3% fathers and) who participated in all follow-up visits had completed at least secondary- or higher-secondary schooling. BL, FU1 and FU2 samples were almost equally distributed across the SES quintiles created at enrolment, with a slightly lower proportion of children from the wealthiest quintile (Q5) being followed-up. Mean height-for-age z-score HAZ increased from -1.57 (SD: 1) at BL, to -1.08 (SD: 1) at FU1 and -0.47 (SD: 0.9) at FU2, and prevalence of stunting reduced from 32.2% at BL to only 6% at FU2.

**Table 2:** Socio-demographic profile of study participants at baseline and follow-up visits 1 and 2.

| **Characteristic** | **Enrolment**<br>**(N=7015)** | **Baseline**<br>**(N=1359)** | **Follow-up 1**<br>**(N=1234)** | **Follow-up 2**<br>**(N=600)** |
| --- | --- | --- | --- | --- |
| Female, n (%) | 3197(45.6) | 623(45.9) | 565(45.8) | 271 (45.2) |
| Age (months), mean (SD) | NA | 39 (1) | 60 (5) | 95 (4) |
| Mother's age at delivery, mean (SD) | (N=6811)<br>23 (4) | 22 (4) | 22 (4) | 22 (4) |
| Mother's education level, n (%) | (N=7010) |  |  |  |
| Below primary (including never been to school) | 807(11.5) | 168(12.4) | 143(11.5) | 71(11.8) |
| Primary/middle school completed | 1754(25) | 350(25.9) | 320(26) | 175(29.2) |
| Secondary/higher secondary school completed | 2591(37) | 525(38.6) | 484(39.2) | 218(36.3) |
| College & above | 1858(26.5) | 316(23.3) | 287(23.3) | 136(22.7) |
| Father's education level, n (%) | (N=7012) |  |  |  |
| Below primary (including never been to school) | 327(4.7) | 72(5.9) | 64(5.2) | 36(6) |
| Primary/middle school completed | 1265(18) | 268(19.7) | 236(19.1) | 127(21.2) |
| Secondary/higher secondary school completed | 3223(46) | 613(45.1) | 566(45.9) | 257(42.8) |
| College & above | 2197(31.3) | 406(29.9) | 368(29.8) | 180(30) |
| SES quintile, n (%)* |  |  |  |  |
| Q1 (poorest) | 1405(20) | 282(20.8) | 256(20.7) | 131(21.8) |
| Q2 | 1403(20) | 306(22.5) | 276(22.4) | 128(21.3) |
| Q3 | 1402(20) | 273(20) | 251(20.3) | 126(21) |
| Q4 | 1403(20) | 264(19.4) | 238(19.3) | 115(19.2) |
| Q5 (wealthiest) | 1402(20) | 234(17.2) | 213(17.3) | 100(16.7) |
| Height-for-age (z-score), mean (SD)* | NA | -1.57 (1) | -1.08(1) | -0.47 (0.9) |
| Stunted, n (%) |  | 437 (32.2) | 214(17.3) | 36(6) |
| School enrolment, n (%)* | NA |  |  |  |
| Private preschools/Schools |  | 329(24.2) | 911(73.8) | 395(65.8) |
| Anganwadi centres |  | 261(19.2) | 63(5.2) | 0(0) |
| Government preschools/ Schools |  | 17 (1.3) | 155(12.5) | 205(34.2) |
| None |  | 752(55.3) | 105(8.5) | 0 |
| BSID-III cognitive domain score, mean (SD) | NA | (N=200) | NA | NA |
|  |  | 69 (5) |  |  |
| CPM Set A, mean (SD) | NA | NA | NA | (N=600)<br>7.75(1.45) |
| CPM Set Ab, mean (SD) | NA | NA | NA | 5.45(1.88) |
| CPM Set B, mean (SD) | NA | NA | NA | 4.65(1.75) |
| CPM Total, mean (SD) | NA | NA | NA | 78.68(11.41) |
| DEEP-score, mean (SD) | NA | 42.39 (5.34) | 58.24 (6.28) | 70.22 (4.88) |
| DEEP-score (Boys), mean (SD) | NA | 42.42 (5.26) | 58.24 (6.10) | 70.21 (4.89) |
| DEEP-score (Girls), mean (SD) | NA | 42.13 (5.44) | 58.23 (6.49) | 70.23 (4.87) |

\* p<0.05

### Criterion Validity

#### Age

The mean DEEP-score was 42.39 (SD: 5.34) at BL when children were 3-years old, 58.24 (SD: 6.28) in 5-year-olds (FU1) and 70.22 (SD: 4.88) in 8-year-olds (FU2) (n=600) and did not differ between boys and girls (Table 2). Pearson’s correlation between DEEP-score and age ranging from 2.5-8-years was 0.87 (CI=0.86-0.89, n=3193) (Figure 2).

**Figure 2:**
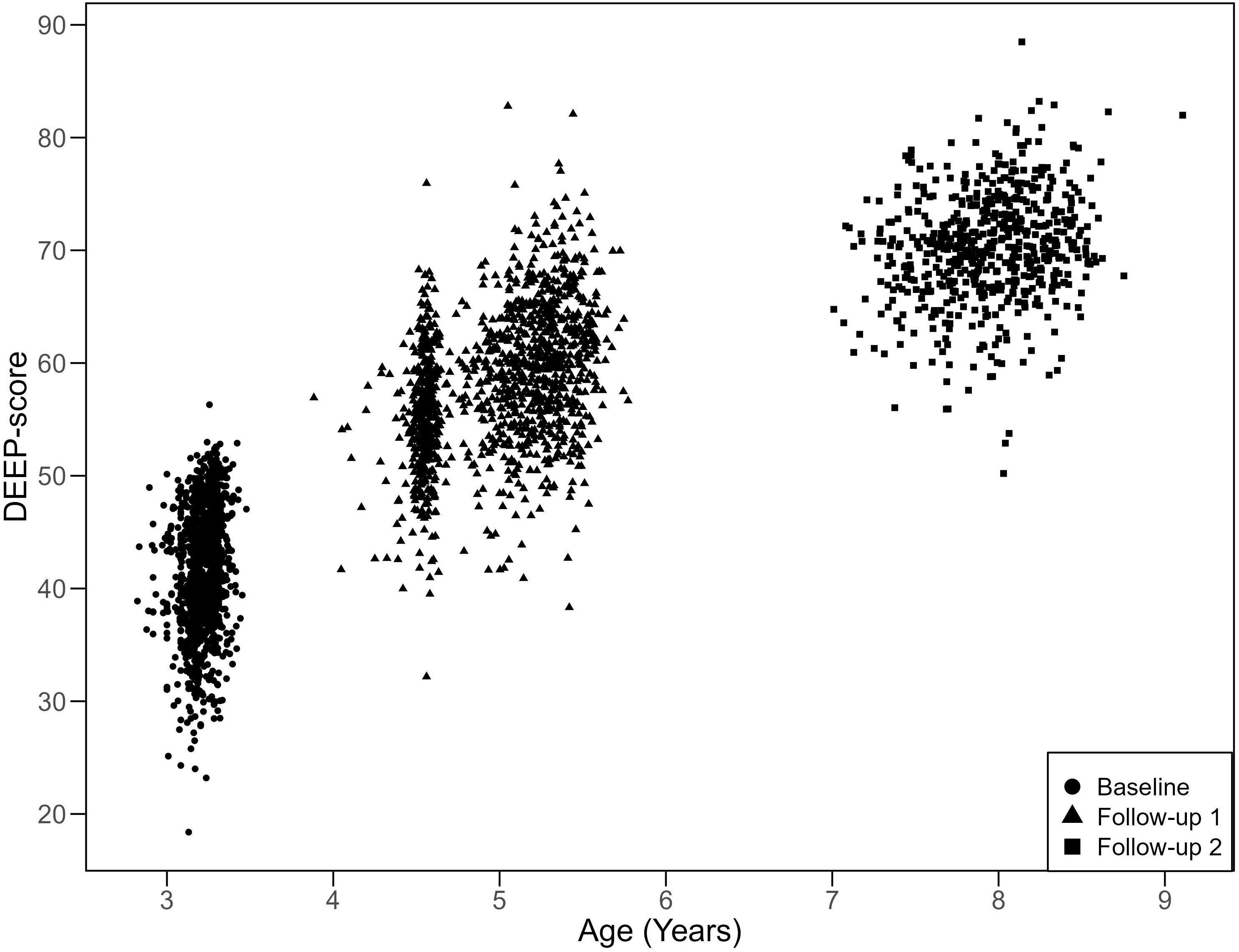
DEEP-score correlates with age across 2.5-8-years (N=3193; r=0.87, CI=0.86 - 0.89). DEEP-score of older children (FU2, 8-year olds – squares) was predicted using the model created on data from preschool-aged children (BL, 3-year olds – circles; FU1, 5-year olds – triangles).

#### BSID-III cognitive domain

DEEP-score was moderately correlated with the cognitive domain score of the BSID-III, a gold-standard clinical assessment, which was concurrently administered on a subset of 200 children when they were 3-years-old (r=0.50, CI:=0.39-0.60) (Table 3). This association was lower than the correlation between the DEEP-score derived using ML (0.67, CI:0.59 - 0.74) (Supplementary Table 3).^17^

**Table 3:**
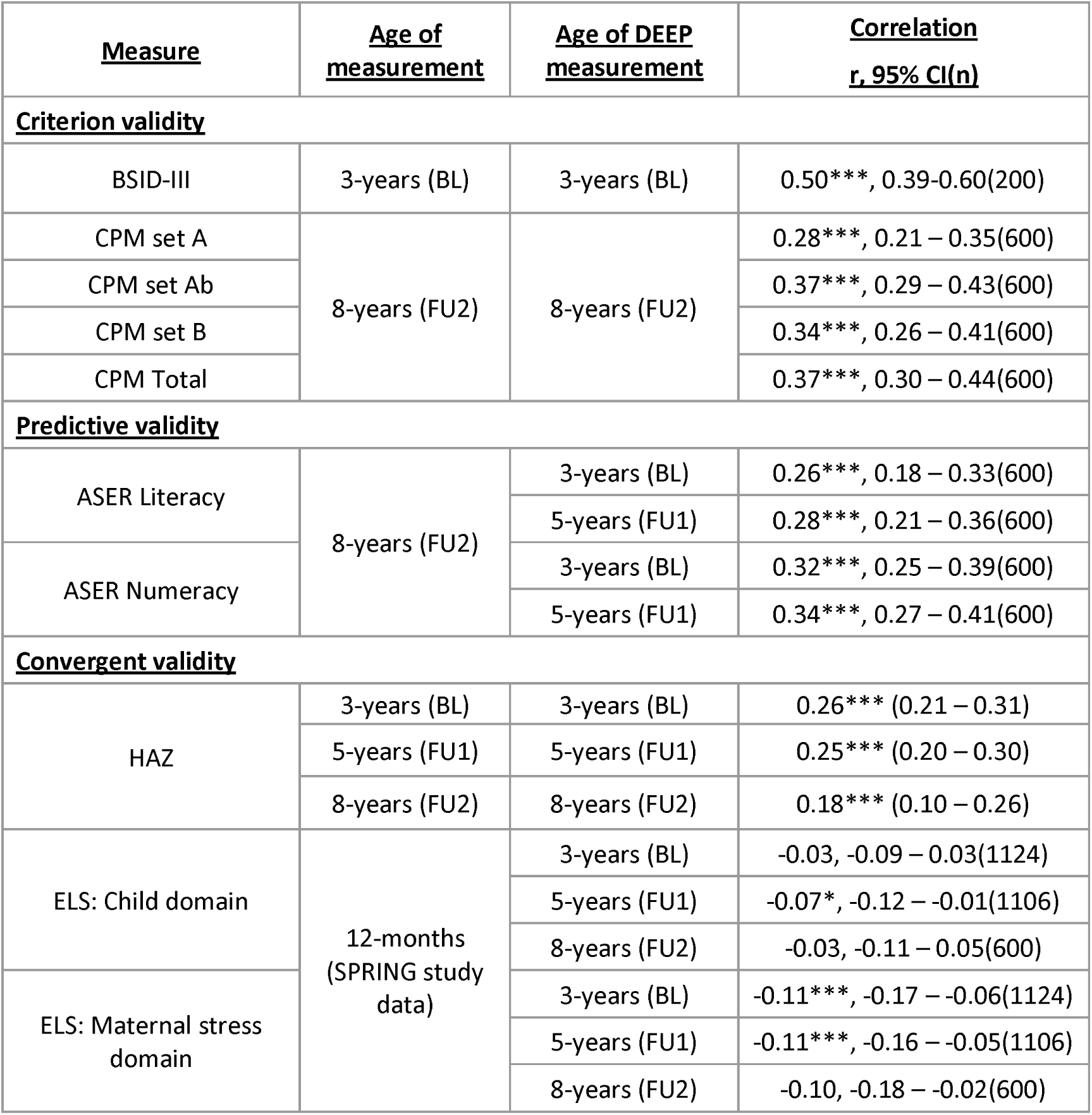
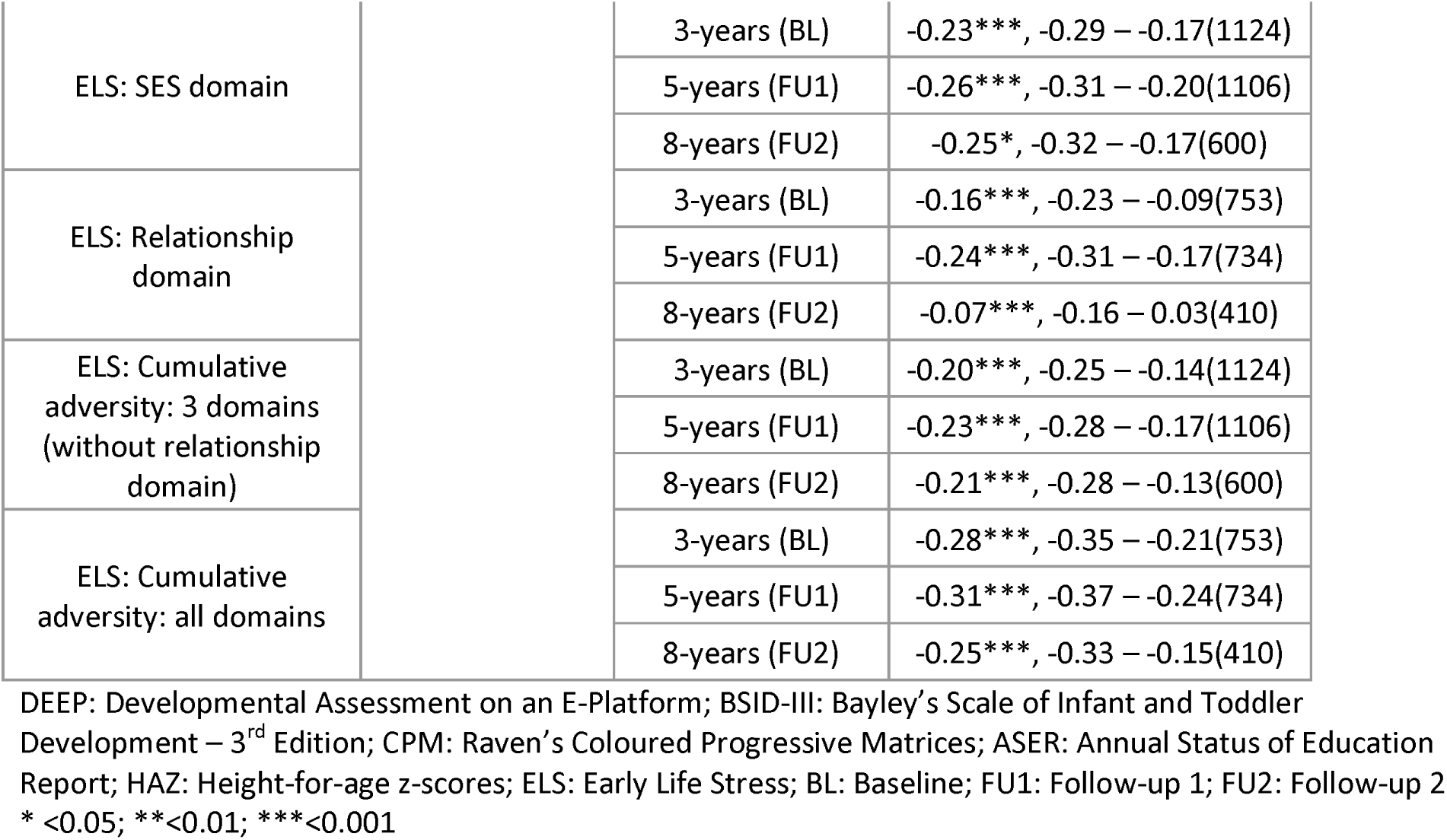
Criterion, predictive and convergent validity of DEEP.

#### CPM

The correlation of DEEP-score of 8-year-old children with total score on concurrently administered assessment fluid intelligence measured using CPM was moderate (0.37; CI: 0.30 – 0.44) and ranged from 0.28 (CI: 0.21 – 0.35) for Set A, 0.34 (CI: 0.26 – 0.41) for Set B and 0.37 (CI: 0.29 – 0.43) for Set Ab (Table 3).

#### Predictive validity

DEEP-score of children in the preschool years (both 3-years and 5-years) predicted their performance on ASER, which measures literacy and numeracy at school-age, when they were 8-years-old (Pearson’s correlation coefficient ranged from 0.26 to 0.35, Table 3), similar to that observed for ML-derived DEEP-score (Supplementary Table 3).

#### Convergent validity

DEEP-score of children at each follow-up was compared with two factors known to relate to cognitive development (Table 3), child linear growth and early-life adversities.

#### Child linear growth

Concurrently measured height-for-age z-scores (HAZ) demonstrated a weak positive correlation with DEEP-score at BL (0.26, CI: 0.21 – 0.31), FU1 (0.25, CI:0.20 – 0.30) and FU2 (0.18, CI:0.10 – 0.26).

#### Early life adversities

Cumulative exposure to adversities in early life when children were 1-year-old correlated negatively with DEEP-score at BL (-0.20; CI:-0.25 – -0.14), FU1 (-0.23; CI:-0.28 – -0.17) and FU2 (-0.21, CI:-0.28 – -0.13) (Table 3), with the association between cognitive development and SES domain being the strongest.

The magnitude of associations of convergent measures with DEEP-score was found to be comparable to their correlations with ASER (Supplementary Table 3), and CPM (Supplementary Table 4).

## Discussion

This study describes the Developmental Assessment on an E-Platform (DEEP) tool and its scoring mechanism using item response theory. DEEP-score is validated through comparisons with a gold-standard clinical cognitive assessment and measures of literacy, numeracy and fluid intelligence. Crucially, the ability of DEEP assessments in the preschool years to predict academic performance in school-age is also demonstrated. Finally, associations between factors known to relate to cognitive development, and DEEP-scores, have been described. To our knowledge, this is the first published study demonstrating the criterion, predictive and convergent validity of a novel scalable digital assessment of cognitive development for preschool children in a large population-based sample from a low-resourced setting, addressing a limitation which has been highlighted recently in the literature.^27^

Participants in this study were recruited at birth and have been followed-up and characterised regularly through the first decade of their life i.e. from birth till middle-childhood. Apart from a slightly lower proportion of families from the wealthiest quintiles participating in these follow-up visits, no significant differences were observed in their socio-demographic profile when compared with the cohort enrolled into the SPRING study, indicating the generalisability of these results. The proportion of children attending formal schooling increased across age with all older children attending school. A drastic reduction was observed in height-for-age z-scores (HAZ) over time in this cohort from 32.2% in 3-year-old children to 6% in 8-year-olds, indicating a high prevalence of catch-up growth in these low-resourced settings within India, similar to findings from an urban poor cohort from Vellore in South India.^28^

To our knowledge, this is the first published study in which item response theory has been used to derive a score of cognitive abilities for preschool children using a combination of *metrics* of child performance recorded by a digital assessment tool. The final model chosen included Accuracy and Completion_time which are most commonly reported for other digital tools.^27^ DEEP-score demonstrates positive correlations with concurrently administered BSID-III, albeit of a lower magnitude than previously published ML-derived DEEP-score which is expected given it was optimised to predict this measure^17^, but still larger than associations demonstrated between BSID-III and other tests like Ages and Stages Questionnaire - 3 (ASQ-3).^29^ Using IRT to score DEEP has the advantage of not relying on being benchmarked to any clinical gold-standard assessments and instead being based on the latent trait of cognitive ability. Another key advantage of using long established IRT methods for score creation over arguably less transparent machine learning methods lies in the rich information, in the form of discrimination and difficulty, IRT provides for every item in the tool allowing for insights into how they are contributing to the tool score allowing for optimisation of its administration and scoring in a data-driven manner in the future as DEEP data continues to be collected in diverse settings. This scoring method will also allow, in the longer term, the use of adaptive testing, i.e., only asking items which are pertinent to a test-taker, to shorten the duration of assessment^19^, to further improve its acceptability, feasibility and scalability.

The strong positive correlation between DEEP-score and age across the preschool years highlights the potential to draw trajectories of cognitive development for this age. DEEP-score at preschool-age predicts children’s literacy and numeracy at school-age, arguably the most crucial property of any developmental assessment. DEEP’s validity is strengthened through its associations with adversities experienced in the first thousand days of life, which are known to exert a long-lasting influence on health and developmental outcomes throughout the life-course.^6^ Significant negative correlations have been demonstrated with cumulative adversity, in particular the socio-economic domain, which included socio-economic status, parental education and family debt or food insecurity, reiterating the importance of these factors on ensuring that children attain their full developmental potential. These demonstrations of criterion, predictive and convergent validity, in addition to its critical advantage of scalability over traditional parent-report or observation-based cognitive assessments, makes a compelling argument for its use in developmental surveillance programs by lay health workers. This would allow early identification of children faltering in their trajectories and the introduction of timely evidence-based interventions while brains are still plastic and retain the potential to respond to their environment.

The participants described in this study represent the follow-up of a population-based birth cohort allowing for analysis of prospective associations, not only between exposures that relate to cognitive outcomes, but also between cognitive measures at different ages making it possible to provide evidence of the predictive validity of DEEP. A limitation of this cohort is that children are not evenly distributed in age across the preschool years making it difficult to draw reference curves for cognitive development based on this dataset. This limitation will be overcome by applying the methods described here on DEEP data collected through an ongoing study, Scalable Transdiagnostic Early Assessment of Mental Health (STREAM),^30^ in which it has been administered on 1080 children each in New Delhi, India and Blantyre, Malawi purposively sampled in quotas which cover the age-range of the tool. Additionally, evidence for DEEP’s reliability (test-retest reliability), structural (the extent to which the empirical correlation structure of the items matches the theorised structure) and cross-cultural validity across diverse settings are not presented here which will also be addressed through the data collected on the STREAM study.

## Data Availability

All data produced in the present study are available upon reasonable request to the authors

## Author contributions

SB, DM, GD, and VP were responsible for the conception and design of the study. SB, AP, AY, CL, KKS, DG, HI, SDT, and DM were responsible for the acquisition and management of data. SB, AR, and GM analysed and interpreted the data. SB and AR drafted the manuscript. GM, and VP reviewed all drafts. All authors reviewed and approved the final version of the manuscript.

## Acknowledgements

The authors would like to thank the SPRING, REACH and COINCIDE consortia and participating families for their support. DEEP was developed in collaboration with the Public Health Foundation of India.

