## Supplementary Material for "A non-specialist worker delivered digital assessment of cognitive development (DEEP) in young children: a longitudinal validation study in rural India"

### **Detailed description of metrics derived from the DEEP tool.**

a) Accuracy: Computed by dividing the number of “correct” clicks or drags from the total number of clicks or drags within each game level, ranges from 0-1.
b) Highest_level: Most games have levels in which the difficulty of the game increases to capture variability in cognitive abilities (see Table 1 for a description of how difficulty is increased in each game). If a child does not successfully complete any difficulty level, the tool automatically terminates the game and presents the child with the next game. Odd One Out (OOO), Series Completion (SC) and Sorting Objects (SO) are designed to measure children’s ability in different dimensions like colour, size and numeracy and each dimension has 3 levels of difficulty. In these games, even if a child failed a difficulty level for one dimension (e.g. colour), they were presented with the next dimension (e.g. size). Highest_Level captures the number of difficulty levels played for each game and can range from 0-15. Exceptions for which this metric was not included are Location Recall (LR) which is woven into the storyline and so the presentation of game levels is not dependent on a child’s ability; Single Tap (ST) and Alternate Tap (AT) which only have 1 difficulty level; Popping Bubbles (PB) in which the presentation of the second difficulty level was not contingent on child performance on the first level.
c) Completion_time: Most games have time limits for completion of each level (see Table 1) such that if a child successfully completes the level within the time limit, the time of the last correct click or drag is recorded, the game level ends and the next difficulty level is presented while if they are unsuccessful within the set time limit, the tool does not present them with the next level but instead proceeds to the next game. Completion_time is the proportion of time within which the child completed the level, derived using the time of the last correct click divided by the time limit for that level, and ranges from 0-1. Exceptions for which this metric was not included are ST, AT and PB the child is presented with a screen on which they play for a fixed period of time; and Grow Your Garden (GYG) since the order in which target and distractor stimuli were presented was randomised implying that it could take longer to complete a level just due to the presentation of stimuli.
d) Latency: Represents the time taken for the first click or drag (could be either correct or incorrect) done by the child in every game level and ranges from 0 to the time limit of each game level.
e) Activity: Computed by dividing the total number of clicks or drags in a game level by Completion_time, except in ST, AT and PB in which they were divided by the game time limit. It represents the number of clicks or drags done by the child per second and ranges from 0.01 to 50.

### **Details of the item-response theory modelling**

The Graded Response Model (GRM) is a widely used item response theory model designed for ordered polytomous response categories. The GRM assumes that each item response can be described by a series of cumulative probabilities corresponding to different difficulty thresholds, which define the boundaries between adjacent response categories. The probability of obtaining $x_{j}$, a given difficulty threshold for item $j$, or higher is given by:

$$P_{x_{j}}^{*}\left( \theta\right)=\frac{e^{\alpha_{j}\left( \theta-\delta_{x_{j}} \right)}}{1+e^{\alpha_{j}\left( \theta-\delta_{x_{j}} \right)}}$$

Where $\theta$ is ability, $\alpha_{j}$ is the discrimination for item $j$, and $\delta_{x_{j}}$ is the difficulty location or threshold for category $x$ for item $j$. Discrimination indicates and item’s ability to differentiate between individuals with different levels of ability. Functionally, discrimination parameters that are too low indicate the item is not useful for measurement of the ability trait while those that are too high (a rare account) indicate the item is only measuring a very specific region of the ability trait. Difficulty indicates the level of ability required to have a 50% chance of answering with a given ordinal category or higher. In practice, we want to use a range of items whose difficulty locations cover the range of abilities we expect to measure.

Data from four of the five metrics, Accuracy, Completion_time, Latency and Activity, was continuous in nature and non-normally distributed. Initially, mixed effect generalised linear regression models were intended to be used to model the scores. However, due to the complexity of jointly modelling the differing implied error distributions (i.e., proportions, time to event), it was decided to segment the metrics so each metric would be distributed as ordinal and IRT analysis could be used to accommodate all metrics. For the metrics Accuracy and Completion_time, data was categorized into equal intervals e.g.., >=0 to <=0.33, >0.33 to <=0.66, and >0.66 to <=1. For Latency and Activity, data was divided into categories, based on terciles in the whole sample. Segmenting the data into 5 categories was not found to improve performance of models derived from single variables and so all subsequent modelling was done using 3 categories (Supplementary Table 1). Further, models using only Latency and Activity data demonstrated poor prediction and were thus excluded from further analysis, and various combinations of the remaining three variables were tested. The final model chosen used three metrics: Accuracy, Highest_level and Completion_time.

The discrimination of the metrics of every game, averaged across game levels, and the difficulty for each game level, averaged across response options, were computed (Supplementary Table 2). The Highest_level metrics for all games and Accuracy and Completion_time for MS, JIG, SO and SD games show high discriminations. Overall, a trend in average game difficulty is observed such that the games which appear first in the tool are easier than those that appear later. However, items did not uniformly increase in difficulty across levels in all games. Test reliability was >0.90 between DEEP scores of 5-75 (Supplementary Figure S1A). The standard error of estimation (SEE) demonstrated the effect of the additional games on increasing the precision of the tool which can be seen when we compare the circles (younger) to the triangles (older) samples (Supplementary Figure S1B).

### **Supplementary Table 1: Models explored to derive DEEP-score.**

Variable combinations used, number of categories into which continuous data was segmented, correlation with age and model fit indices across explored models. The final chosen model (S.No. 10) is in bold.

| **S.No.** | **Model** | **Variables** | **Categories** | **Correlation with age**  **r (95% CI)** | **RMSEA** | **TLI** | **CFI** |
| --- | --- | --- | --- | --- | --- | --- | --- |
| Single variables | | | | | | | |
| 1 | Highest_level | 1 | 0-15 | 0.87 (0.86 – 0.89) *** | 0.04 | 0.99 | 1.00 |
| 2 | Accuracy | 1 | 3 | 0.78 (0.76 – 0.79) *** | 0.02 | 0.99 | 0.99 |
| 3 | Accuracy |  | 5 | 0.79 (0.78 – 0.80) *** | 0.03 | 0.99 | 0.99 |
| 4 | Completion_time | 1 | 3 | 0.69 (0.67 – 0.71) *** | 0.02 | 1.00 | 1.00 |
| 5 | Completion_time |  | 5 | 0.70 (0.68 – 0.72) *** | 0.02 | 0.99 | 0.99 |
| 6 | Latency | 1 | 3 | 0.08 (0.04 – 0.11) *** | 0.03 | 0.88 | 0.88 |
| 7 | Latency |  | 5 | 0.03 (-0.01 – 0.07) **ns** | 0.04 | 0.9 | 0.9 |
| 8 | Activity | 1 | 3 | 0.13 (0.09 – 0.17) *** | 0.04 | 0.91 | 0.91 |
| 9 | Activity |  | 5 | 0.23 (0.19 – 0.27) *** | 0.05 | 0.91 | 0.91 |
| Combining variables | | | | | | | |
| **10** | **Highest_level + Accuracy + Completion_time** | **3** | **3** | **0.83 (0.82 – 0.84) ***** | **0.03** | **0.99** | **0.99** |
| 11 | Accuracy + Completion_time | 2 | 3 | 0.78 (0.76 – 0.79) *** | 0.03 | 0.99 | 0.99 |
| 12 | Highest_level + Accuracy | 2 | 3 | 0.85 (0.84 – 0.86) *** | 0.03 | 0.99 | 0.99 |
| 13 | Highest_level + Completion_time | 2 | 3 | 0.82 (0.81 – 0.83) *** | 0.02 | 0.99 | 0.99 |

RMSEA = root mean square error of approximation, TLI = Tucker-Lewis Index, CFI = Comparative Fit Index (CFI); *<0.05; **<0.01; ***<0.001; ns ≥ 0.05

### **Supplementary Table 2: Discrimination and difficulty of DEEP items**

The discrimination of the metrics of every game, averaged across game levels, and the difficulty for each game level, averaged across response options (grey colour = not applicable) for the final chosen model.

|  | **Game** | **Avg. Discrimination** | **Avg. Difficulty** | **L1** | **L2** | **L3** | **L4** | **L5** | **L6** | **L7** | **L8** | **L9** | **L10** | **L11** | **L12** | **L13** | **L14** | **L15** |
| --- | --- | --- | --- | --- | --- | --- | --- | --- | --- | --- | --- | --- | --- | --- | --- | --- | --- | --- |
| **Highest_level** | LR |  |  |  |  |  |  |  |  |  |  |  |  |  |  |  |  |  |
|  | ST |  |  |  |  |  |  |  |  |  |  |  |  |  |  |  |  |  |
|  | AT |  |  |  |  |  |  |  |  |  |  |  |  |  |  |  |  |  |
|  | PB |  |  |  |  |  |  |  |  |  |  |  |  |  |  |  |  |  |
|  | GYG | 1.73 | -0.89 |  |  |  |  |  |  |  |  |  |  |  |  |  |  |  |
|  | HO | 2.54 | -1.20 |  |  |  |  |  |  |  |  |  |  |  |  |  |  |  |
|  | OOO | 3.10 | -1.04 |  |  |  |  |  |  |  |  |  |  |  |  |  |  |  |
|  | SD | 2.28 | -0.89 |  |  |  |  |  |  |  |  |  |  |  |  |  |  |  |
|  | MS | 5.06 | -0.30 |  |  |  |  |  |  |  |  |  |  |  |  |  |  |  |
|  | JIG | 4.12 | -0.39 |  |  |  |  |  |  |  |  |  |  |  |  |  |  |  |
|  | SO | 3.34 | -0.54 |  |  |  |  |  |  |  |  |  |  |  |  |  |  |  |
|  | SC | 2.86 | -0.07 |  |  |  |  |  |  |  |  |  |  |  |  |  |  |  |
|  | PM | 2.61 | 0.84 |  |  |  |  |  |  |  |  |  |  |  |  |  |  |  |
|  | SR | 2.03 | 0.48 |  |  |  |  |  |  |  |  |  |  |  |  |  |  |  |
| **Accuracy** | LR | 0.91 | -1.13 | -1.84 | -1.64 | -1.12 | -0.88 | -0.22 | -0.95 | -1.18 | -1.24 |  |  |  |  |  |  |  |
|  | ST | 0.89 | -3.99 | -3.99 |  |  |  |  |  |  |  |  |  |  |  |  |  |  |
|  | AT | 2.26 | -1.40 | -1.40 |  |  |  |  |  |  |  |  |  |  |  |  |  |  |
|  | PB | 1.58 | -1.78 | -1.90 | -1.67 |  |  |  |  |  |  |  |  |  |  |  |  |  |
|  | GYG | 1.29 | -1.50 | -2.10 | -1.76 | -2.29 | 1.12 | -2.51 |  |  |  |  |  |  |  |  |  |  |
|  | HO | 1.42 | -1.59 | -2.16 | -1.38 | -0.70 | -0.73 | -0.61 | -3.17 | -2.37 |  |  |  |  |  |  |  |  |
|  | OOO | 0.81 | -0.13 | -0.68 | -1.27 | -0.62 | -0.13 | 0.15 | -0.43 | -0.93 | -0.56 | 0.58 | -1.46 | 1.51 | 0.88 | 0.33 | 0.03 | 0.67 |
|  | SD | 1.49 | -0.22 | -0.57 | 0.08 | -0.15 |  |  |  |  |  |  |  |  |  |  |  |  |
|  | MS | 2.16 | -0.40 | -0.51 | -0.82 | -0.87 | -0.42 | -0.25 | -0.43 | 0.75 | -0.36 | -0.69 |  |  |  |  |  |  |
|  | JIG | 1.59 | -0.33 | -0.85 | -1.12 | -0.69 | -0.49 | -0.48 | -0.56 | 0.56 | 0.69 | -0.01 |  |  |  |  |  |  |
|  | SO | 1.48 | -0.98 | -2.86 | -0.46 | -0.73 | -0.32 | -1.07 | -1.30 | -0.72 | -0.44 | -0.87 |  |  |  |  |  |  |
|  | SC | 0.91 | 0.46 | 1.24 | 0.69 | 1.09 | 0.68 | 0.75 | -2.56 | 1.21 | 0.06 | 0.99 |  |  |  |  |  |  |
|  | PM | 0.76 | 0.48 | 0.85 | -2.33 | 2.92 |  |  |  |  |  |  |  |  |  |  |  |  |
|  | SR | 0.94 | 1.12 | -0.58 | 1.08 | 2.86 |  |  |  |  |  |  |  |  |  |  |  |  |
| **Completion_time** | LR | 1.15 | -4.11 | -6.37 | -5.08 | -4.15 | -3.12 | -3.28 | -3.62 | -3.57 | -3.72 |  |  |  |  |  |  |  |
|  | ST |  |  |  |  |  |  |  |  |  |  |  |  |  |  |  |  |  |
|  | AT |  |  |  |  |  |  |  |  |  |  |  |  |  |  |  |  |  |
|  | PB |  |  |  |  |  |  |  |  |  |  |  |  |  |  |  |  |  |
|  | GYG |  |  |  |  |  |  |  |  |  |  |  |  |  |  |  |  |  |
|  | HO | 1.06 | -1.50 | -3.52 | -2.33 | -1.26 | -1.59 | -1.02 | -1.28 | 0.48 |  |  |  |  |  |  |  |  |
|  | OOO | 1.08 | -4.07 | -2.31 | -2.71 | -3.90 | -1.92 | -1.09 | -2.65 | -2.45 | -2.06 | -29.17 | -3.61 | -3.53 | -1.99 | -0.89 | -1.45 | -1.42 |
|  | SD | 1.98 | -0.97 | -1.31 | -0.48 | -1.12 |  |  |  |  |  |  |  |  |  |  |  |  |
|  | MS | 3.11 | -0.47 | -0.70 | -0.39 | -1.01 | -0.73 | -0.57 | -0.31 | 0.07 | -0.29 | -0.32 |  |  |  |  |  |  |
|  | JIG | 2.07 | -0.34 | -0.94 | -1.13 | -0.91 | -1.05 | -0.66 | -0.51 | 1.02 | 0.78 | 0.32 |  |  |  |  |  |  |
|  | SO | 2.50 | -0.65 | -1.12 | -0.39 | -0.54 | -0.22 | -0.81 | -0.96 | -0.83 | -0.34 | -0.62 |  |  |  |  |  |  |
|  | SC | 1.18 | -0.64 | 0.96 | 0.31 | -0.32 | 0.39 | -0.09 | -2.99 | 0.29 | -2.05 | -2.27 |  |  |  |  |  |  |
|  | PM | 0.93 | 0.22 | 0.86 | 1.52 | -1.73 |  |  |  |  |  |  |  |  |  |  |  |  |
|  | SR | 1.37 | -0.85 | -1.64 | -0.45 | -0.46 |  |  |  |  |  |  |  |  |  |  |  |  |

### **Supplementary Table 3: Correlations between Annual Status of Education Report (ASER) and measures used for DEEP’s convergent validity.**

Associations of DEEP-score, ASER Literacy and ASER Numeracy with HAZ and early life adversities.

| **Measure, age** | **DEEP-score r, 95% CI (n)** | **ASER Literacy r, 95% CI (n)** | **ASER Numeracy r, 95% CI (n)** |
| --- | --- | --- | --- |
| Height-for-age z score (HAZ), 8-years (FU 2) | 0.18***, 0.10 - 0.26 (600) | 0.21***, 0.13 - 0.28 (600) | 0.25***, 0.18 - 0.33 (600) |
| Child domain, 12-months (SPRING study data) | -0.03, -0.11 - 0.05 (600) | -0.08*, -0.16 - 0.00 (600) | -0.12**, -0.19 - -0.04 (600) |
| Maternal stress domain, 12-months (SPRING study data) | -0.10, -0.18 - -0.02 (600) | -0.03, -0.13 - 0.06 (410) | -0.11*, -0.21 - -0.01 (410) |
| SES domain, 12-months (SPRING study data) | -0.25*, -0.32 - -0.17 (600) | -0.13**, -0.21 - -0.05 (600) | -0.14***, -0.22 - -0.06 (600) |
| Relationship domain, 12-months (SPRING study data) | -0.07***, -0.16 - 0.03 (410) | -0.33***, -0.40 - -0.26 (600) | -0.35***, -0.42 - -0.28 (600) |
| Cumulative adversity: 3 domains (without relationship domain), 12-months (SPRING study data) | -0.21***, -0.28 - -0.13 (600) | -0.29***, -0.36 - -0.21 (600) | -0.32***, -0.39 - -0.25 (600) |
| Cumulative adversity: all domains, 12-months (SPRING study data) | -0.25***, -0.33 - -0.15 (410) | -0.30***, -0.39 - -0.21 (410) | -0.36***, -0.44 - -0.28 (410) |

*<0.05; **<0.01; ***<0.001

### **Supplementary Table 4: Correlations between Raven’s Coloured Progressive Matrices (CPM) and measures used for DEEP’s convergent validity.**

Associations of DEEP-score and CPM scores with HAZ and early life adversities.

| **Measure, age** | **DEEP-score r, 95% CI (n)** | **CPM set A r, 95% CI (n)** | **CPM set Ab r, 95% CI (n)** | **CPM set B r, 95% CI (n)** | **CPM Total r, 95% CI (n)** |
| --- | --- | --- | --- | --- | --- |
| Height-for-age z score (HAZ), 8-years (FU 2) | 0.18***, 0.10 - 0.26 (600) | 0.14***, 0.06 - 0.21 (599) | 0.14***(0.06 - 0.22 (599) | 0.13**, 0.05 - 0.21 (599) | 0.17***, 0.09 - 0.25 (599) |
| Child domain, 12-months (SPRING study data) | -0.03, -0.11 - 0.05 (600) | -0.03, -0.11 - 0.05 (599) | -0.03, -0.11 - 0.05 (599) | -0.05, -0.13 - 0.03 (599) | -0.05, -0.13 - 0.03 (599) |
| Maternal stress domain, 12-months (SPRING study data) | -0.10, -0.18 - -0.02 (600) | -0.11*, -0.20 - -0.01 (409) | -0.17***, -0.26 - -0.07 (409) | -0.11*, -0.21 - -0.02 (409) | -0.20***, -0.29 - -0.11 (409) |
| SES domain, 12-months (SPRING study data) | -0.25*, -0.32 - -0.17 (600) | -0.09*, -0.17 - -0.01 (599) | -0.09*, -0.17 - -0.01 (599) | -0.06, -0.14 - 0.02 (599) | -0.11**, -0.19 - -0.03 (599) |
| Relationship domain, 12-months (SPRING study data) | -0.07***, -0.16 - 0.03 (410) | -0.12**, -0.20 - -0.04 (599) | -0.15***, -0.23 - -0.08 (599) | -0.18***, -0.26 - -0.10 (599) | -0.21***, -0.28 - -0.13 (599) |
| Cumulative adversity: 3 domains (without relationship domain), 12-months (SPRING study data) | -0.21***, -0.28 - -0.13 (600) | -0.12**, -0.20 - -0.05 (600) | -0.14***, -0.22 - -0.06 (600) | -0.16***, -0.23 - -0.08 (600) | -0.20***, -0.27 - -0.12 (599) |
| Cumulative adversity: all domains, 12-months (SPRING study data) | -0.25***, -0.33 - -0.15 (410) | -0.16**, -0.25 - -0.07 (409) | -0.22***, -0.31 - -0.12 (409) | -0.21***, -0.30 - -0.12 (409) | -0.28***, -0.37 - -0.19 (409) |

*<0.05; **<0.01; ***<0.001

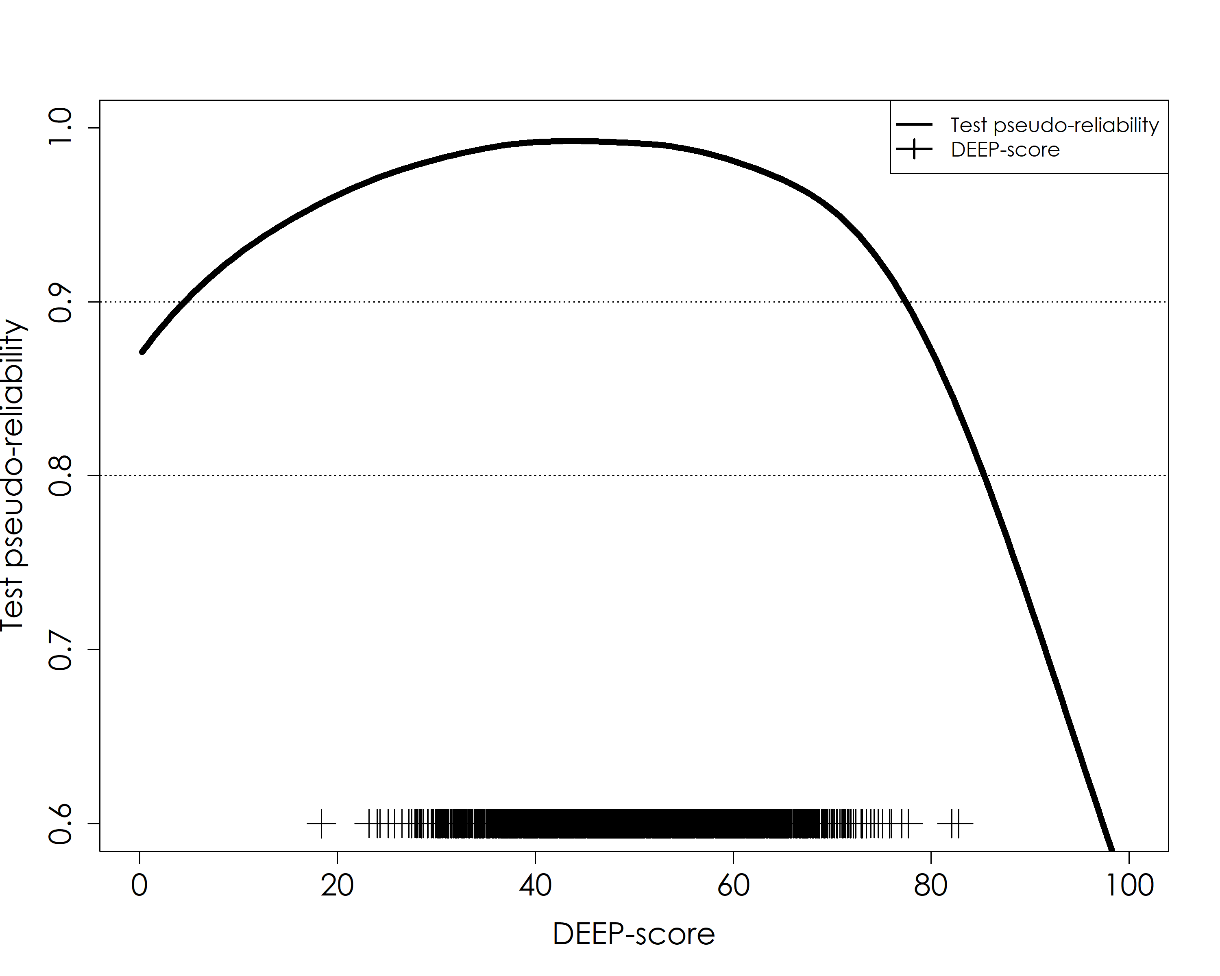

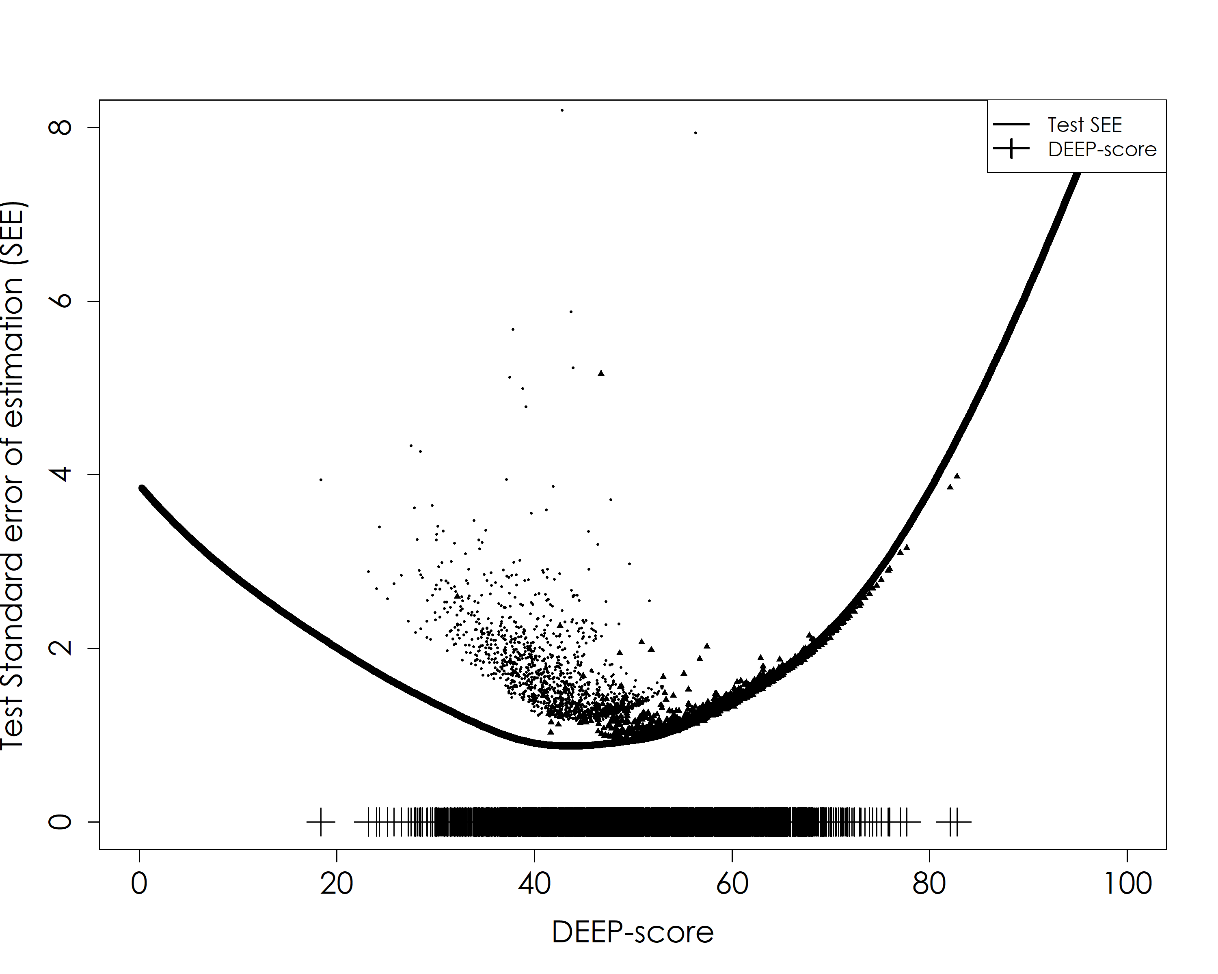

**Supplementary Figure S1: Test pseudo-reliability and standard error of estimation of the final model for DEEP-score.** (A) Test pseudo-reliability of DEEP-scores; (B) The Standard Error of Estimation (SEE) for younger (circles) and older (triangles) children.
